# Somatic mutations in both blood cells and vascular endothelium from patients with severe coronary artery diseases

**DOI:** 10.1101/2022.01.17.22269173

**Authors:** Haotian Zhang, Henry Tannous, Christopher Mazzeo, Haoyi Zheng, Huichun Zhan

## Abstract

Individuals with clonal hematopoiesis of indeterminate potential (CHIP) is associated with a 2-4 fold increase in cardiovascular diseases (CVDs). While most studies focus on mutant blood cells, we propose that a better insight into the CHIP-CVD connection can be obtained by examining the genetic mutations in both the vascular endothelium and peripheral blood samples from the same patient. In this study, we examined somatic mutations by whole exome sequencing in paired blood and vascular samples from 4 patients with severe coronary artery disease who underwent coronary artery bypass graft surgery. We found that blood and endothelium can acquire the same mutations during aging and CHIP-associated mutations are not uncommon in vascular endothelium in patients with severe CVDs. Since vascular endothelial cells have critical roles in the regulation of cardiovascular function, mutant vascular endothelium offers an opportunity for further investigations to improve our diagnosis and management of individuals with CHIP.

---

In recent years, it has been recognized that expansion of mutated blood cells in individuals without frank malignancy, or clonal hematopoiesis of indeterminate potential (CHIP), is present in 10-20% of people older than 70 yrs^1-3^. CHIP carriers have a 2-4 fold increase in cardiovascular diseases (CVDs) with worsened clinical outcomes, which rivals or even exceeds those traditional CVD risk factors such as diabetes, hypertension, hyperlipidemia, and smoking^3^. While most studies focus on mutant blood cells, we propose that a better insight into the CHIP-CVD connection can be obtained by examining the genetic mutations in both the vascular endothelium and peripheral blood samples from patients with CVDs.

Patients with severe coronary artery diseases (CADs) often undergo coronary artery bypass graft (CABG) surgery during which segments of arterial (e.g., internal mammary artery) or venous (e.g., saphenous vein) grafts are used to bypass the diseased coronary arteries. Excess vascular tissue is available from these surgical procedures. In this study, we performed whole exome sequencing in paired vascular graft and peripheral blood samples from four consented patients (2 females and 2 males, age 68-77yr old) with severe CADs who underwent CABG surgery. Vascular samples were washed to remove any residual blood cells and thinned to remove subcutaneous tissue. For patients #1-#3, genomic DNA was extracted directly from a ∼5mm piece of the vascular tissue (Figure 1A). For patient #4, the vascular tissue was digested to obtain single cells, followed by flow cytometry sorting to obtain CD31^+^CD45^-^ endothelial cells (ECs) and genomic DNA was extracted from the isolated ECs. The CD31^+^CD45^-^ cells were also positive for CD34 and CD144 (not shown) and their EC identity was confirmed using both tube formation assay (as a measure of angiogenesis *in vitro*) and immunofluorescence staining (for EC-specific markers von Willebrand Factor and CD31) (Figure 1B).

**Figure 1.**
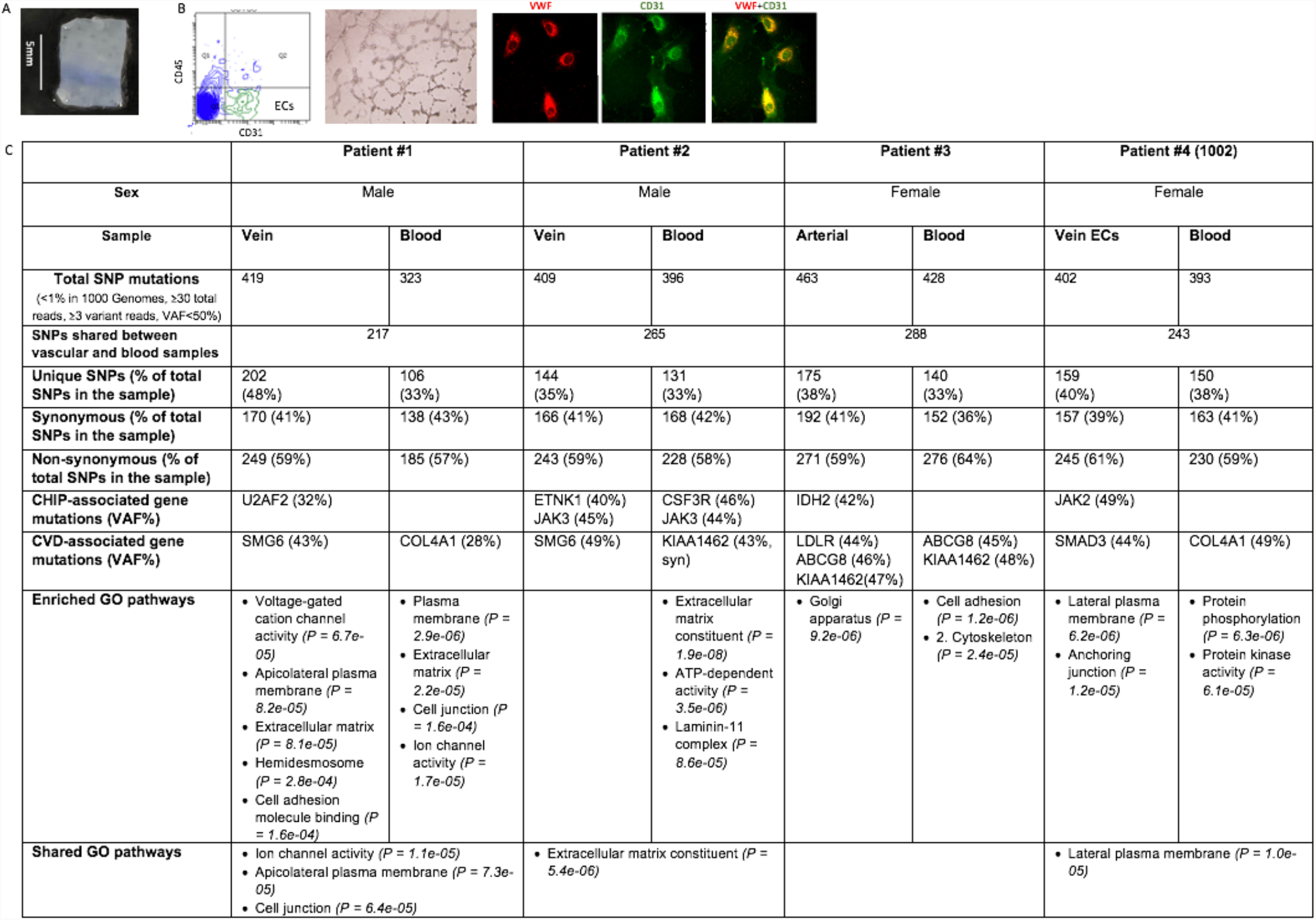
SNP mutations assayed from 4 patients with severe coronary artery diseases. (**A**) A representative cleaned and excised vascular sample from patients #1-#3. (**B**) (*left*) Flow cytometry plot depicting the gating strategy used to sort CD45^-^CD31^+^ ECs from the vascular sample of patient #4. (*middle*) Sorted ECs were seeded in Matrigel and tube formation was observed after an 8-hour incubation (magnification: 10x). (*right*) The fluorescence image of ECs with positive Von Willebrand Factor (VWF) (red) and CD31 (green) staining (magnification: 40x). (**C**) SNP mutations detected in paired vascular and blood samples from 4 patients.

Whole exome sequencing data were aligned to the human reference genome hg38 using Burrows-Wheeler Aligner. SAMtools was used for sorting the BAM files, and Picard was utilized to mark duplicate reads. 60-95% of the targets were covered with at least 50x depth. Single nucleotide polymorphisms (SNPs) were identified by GATK. False-positive filters were applied and mutations affecting 3’ or 5’ UTR, intronic or intergenic sequences, or segmental duplications of the human genome were discarded prior to downstream analysis. To limit germline variants and potential artifacts, variants with observed frequency >1% in the 1000 Genomes reference or variant allele fraction (VAF) more than 50% of the sequencing reads arising from that genomic site were excluded^1,2^. Variants were also excluded if the sequencing depth at variant site was <30 and the number of reads supporting variant alleles was <3.

∼400 candidate somatic SNPs were identified in patients #1-#3, and ∼250 of these SNPs were shared between the blood and vascular samples. To make sure this is not due to any blood cell contamination of the vascular sample, we isolated vascular ECs from patient #4 by flow cytometry sorting; whole exome sequencing of the isolated ECs revealed similar results. Gene ontology analysis of all the candidate SNPs revealed that, although different pathways were enriched in the two tissues, pathways involved in plasma membrane, cell junction, and extracellular matrix were shared between the vascular and blood samples, suggesting that interactions between the mutant blood cells and mutant endothelium may play a key role in the development of CVD in these patients. (Figure 1C)

We then set out to identify CHIP carriers on the basis of a pre-specified list of 74 genes known to be recurrently mutated in myeloid cancers^3^. Although CHIP-associated mutations were only detected in 1 blood sample of patient #2 (CSF3R, JAK3), CHIP mutations (JAK2, JAK3, IDH2, U2AF2, ETNK1) were present in all 4 vascular samples with VAFs between 32-49%. We also searched the literature and compiled a list of 90 genes associated with CVDs^4,5^. CVD-associated gene mutations were detected in both the vascular and blood samples of all 4 patients; in particular, SMG6, COL4A1, and KIAA1462 were mutated in 2 of the 4 patients. Although some of these CHIP and CVD-associated mutations were detected in both the blood and vascular samples from the same patient (e.g., JAK3 in patient #2, ABCG8 and KIAA1462 in patient #3), most mutations were unique to either the blood sample or the vascular sample, further indicating that these were neither due to blood cell contamination nor due to germline mutation.

Although a common origin between blood and endothelium beyond the embryonic stage of development continues to be a subject of intense investigation, results from this study suggest that blood and endothelium can acquire the same mutations during aging and CHIP-associated mutations are not uncommon in vascular endothelium in patients with severe CVDs. Mutant vascular endothelium offers an opportunity for further investigations to improve our diagnosis and management of individuals with CHIP.

## Data Availability

All data produced in the present study are available upon reasonable request to the authors

## ACKNOWLEDGEMENTS

This research was supported by the National Heart, Lung, and Blood Institute grant NIH R01 HL134970 (H.Z.), VA Career Development Award BX001559 (HZ), and VA Merit Award BX003947 (H.Z.).

